# A Minimally Invasive, Suturable Platform for Brain Monitoring

**DOI:** 10.64898/2026.06.24.26355847

**Authors:** Abdulhameed Abdal, Hyeonseok Kim, Fadi Khoury, Peter Hadar, Brian F. Coughlin, Paige Schumsky, Gillian Celis, Jason Chin, Anthony Navarro, Sarine Krikorian, Samuel J. Edmunds, Ritwik Vatsyayan, Moumita Halder, Tarek Rafeedi, Jiahao Wan, Rachel Blau, Kaushik Shukla, Tianhai Wu, Jesse Jokerst, Darren J. Lipomi, Sydney S. Cash, Shadi A. Dayeh

## Abstract

Capturing infrequent or context-dependent brain events, such as epileptic seizures and sleep abnormalities, often requires continuous monitoring over several days. It is most practical and scalable when achieved with unobtrusive, high-fidelity wireless systems that patients can use at home. Current electroencephalography systems restrict patient mobility and require continuous electrode maintenance and sub-scalp solutions require surgical implantation that offer limited spatial coverage. We developed NeuroWeaves, gold–polyimide microthreads thinner than a human hair that can be stitched through the epidermis using standard suture tools and connected to a lightweight wireless recorder. In preclinical models, NeuroWeaves captured whisker-evoked potentials with accuracy comparable to skull screws and matched the performance of a commercial acquisition benchmark. Semi-chronic recordings in freely moving animals remained stable for several weeks, and 30-day histology showed minimal inflammation comparable to surgical sutures. Pilot human studies reproduced posterior-dominant rhythms, photic responses, chewing artifacts, and sleep oscillations comparable to clinical electrodes. These results establish a minimally invasive, biocompatible neural interface that represents a new modality for high-fidelity brain monitoring beyond conventional laboratory and clinical constraints.

## Main

Continuous electroencephalography (EEG) is foundational to epilepsy diagnosis^1^, sleep research^2^, and neuro-ergonomics^3^, yet no available electrode system achieves the combined requirements of long-term comfort, signal stability, and high fidelity in real-world settings. Clinical EEG remains reliant on gel-based surface electrodes that can require abrasive skin preparation, conductive paste, and frequent technician intervention^4^. As the gels dry, the interface impedance increases and the signal quality deteriorates, which imposes a substantial maintenance burden for human subjects and clinical staff^5^. Because seizure frequency varies widely across individuals, single-session recordings and patient self-report remain unreliable, and continuous multi-day monitoring is needed for precise characterization^6,7^. Costs further exacerbate this gap. Hospitalization in an epilepsy monitoring unit (EMU) can exceed $40,000 per admission^8^. This cost limits access in regions where ∼80% of the world’s 5 million new epilepsy cases occur each year^9^.

A recently emerging class of “sub-scalp” technologies seeks to address these limitations by placing electrodes in the sub-galeal space, thereby bypassing the high-impedance epidermal barrier^10^. These systems leverage macroscale contacts (∼10 mm width) to achieve sub-kilohm (kΩ) impedance and long-term stability^11,12^. However, their clinical adoption remains constrained: The bulky form factors require centimeter-scale incisions, careful tunneling, and general anesthesia in an operating room, deterring many prospective patients^13,14^. Reimbursement pathways remain unknown, and currently approved devices (e.g., Minder^TM^ and UNEEG SubQ^TM^) offer only unilateral or limited bilateral coverage, restricting spatial sampling^13^. Reported complications—including scalp paresthesia, headaches, and sub-scalp hematoma—further underscore the limitations of current systems^10,14,15^. Collectively, these issues highlight the need for a less invasive platform that supports high-fidelity, fully ambulatory brain monitoring.

We developed NeuroWeaves, a minimally invasive recording modality (**Fig. 1a**) designed to address these barriers through four integrated advances in clinical electrophysiology. First, stitch-based electrodes that anchor within viable epidermis and can be implanted without anesthesia using a workflow identical to routine clinical suturing. Second, a hair-thin (**Fig. 1b**), customizable thread architecture allows modulation of device length, contact spacing, and layout. Third, adaptable implantation configurations can be selected to align with distinct clinical objectives. Finally, a compact wearable wireless system enables continuous raw EEG monitoring outside of traditional hospital infrastructure. These features provide stable recording performance while prioritizing comfort, ease of use, and operational feasibility for long-duration monitoring.

**Fig. 1.**
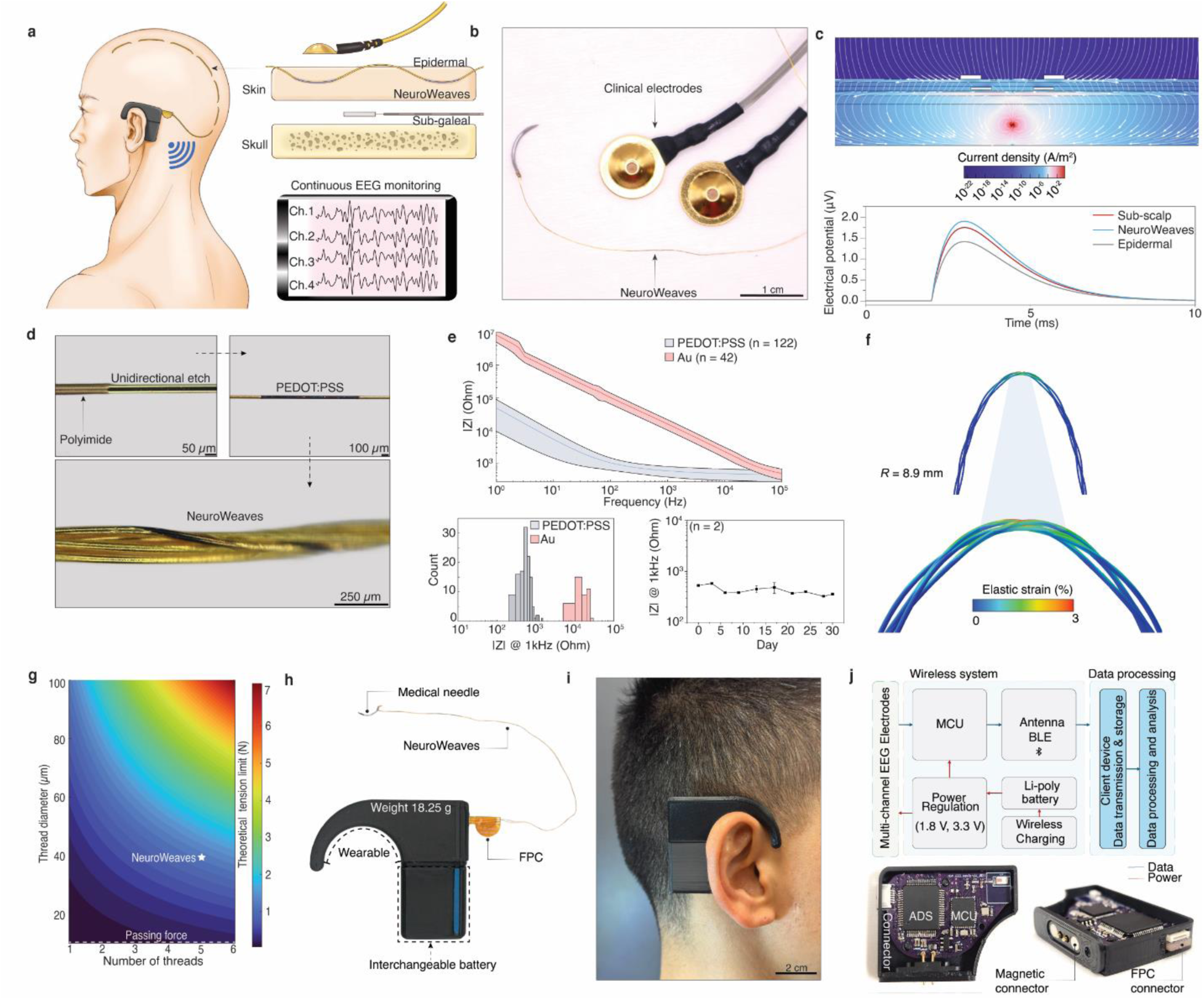
Overview of NeuroWeaves. **a**, Schematic illustration of the wireless integrated NeuroWeaves system. Inset shows comparison of NeuroWeaves to current clinical modalities. **b**, Photographic image of hair-thin NeuroWeaves device compared to clinical scalp electrodes. **c**, Simulation results of current density distribution across modalities (top) and electrical potential is highest for NeuroWeaves at 1.5 cm inter-electrode pitch (bottom). **d**, Photographic images of the fabrication process of gold-polyimide filaments. **e**, Electrochemical impedance before (n = 42) and after (n = 122) electrodeposition of PEDOT:PSS across 1 – 10^5^ frequency spectra (top). Histogram shows at least a magnitude difference between PEDOT:PSS and Au contacts at 1 kHz frequency (bottom, left). In vitro aging test of pristine PEDOT:PSS in 37 °C saline show stability of impedance at 1 kHz frequency across 30 days study (bottom, right). **f**, Simulations show bending up to small curvature of 8.9 mm results an elastic strain of 3%. **g**, Theoretical simulation of thread diameter and number of threads for choice of design. Star is the choice of design of NeuroWeaves for initial tests showing theoretical tension limit ∼ 2 N, which is close to experimental tensile tests of ∼1.3 N. Dashed line resembles passing force of NeuroWeaves we performed to measure interfacial tension when passing through porcine skin. **h**, Photographic image of the integrated wireless wearable system when attached to the NeuroWeaves device. **i**, Photographic image of human subject demonstrating wearability of the cochlear inspired wireless module. **j**, Diagram showing the data and power flow for the wireless module (top) and internal components of the printed-circuit board (PCB) when housed in a compact 3-D printed design with attached magnetic connectors for interchangeable batteries (bottom).

We first characterize the fabrication, mechanical properties, and implantation workflow of NeuroWeaves, followed by validation in acute preclinical recordings using both commercial tethered and custom wireless acquisition systems. We also evaluate the ability of NeuroWeaves to record chemically induced epileptic seizures. Next, we assess semi-chronic performance in freely moving animals and finally demonstrate clinical translation through recordings in a healthy participant. Collectively, these studies evaluate recording performance, stability, and translational feasibility across preclinical and clinical settings.

## Results

### Device Architecture and Wearable Integration of NeuroWeaves

To implement this system, we engineered NeuroWeaves at the level of the device, materials, fabrication, and system integration. Spatial resolution and signal strength are jointly determined by electrode pitch in EEG systems^16,17^. Based on simulations and human-use constraints, we selected a 1.5-cm inter-electrode pitch to maximize amplitude capture at the cutaneous surface (**Fig. 1c**, **Supplementary Fig. 1**, and **Tables 1** to **2**). Each NeuroWeave consisted of an insulated gold thread (∼45 µm in diameter) (**Supplementary Fig. 2**), which was selectively etched to expose a millimeter-scale recording site and then multiple of the threads were twisted together to form the final device structure (**Fig. 1d** and **Supplementary Fig. 3**). Wafer-scale batch processing is amenable to diverse device geometries for preclinical and clinical applications, as well as cost-efficient production (**Supplementary Fig. 4** and **Table 3**). We refined the fabrication scheme by transitioning from multi-step photolithography to a streamlined hard-mask process using thermal release tapes, which reduced fabrication downtime and increased overall throughput (fabrication steps and characterizations are available in **Supplementary Figs. 5** to **8**).

Interfacial impedance is a key determinant of bioelectronic signal quality^18^. Clinical guidelines from the American Clinical Neurophysiology Society (ACNS) recommend electrode impedances between 5 – 10 kΩ for reliable EEG acquisition^19^. Because each NeuroWeaves contact has an exceptionally small geometrical area, achieving low impedance requires a material with high volumetric capacitance and proven biocompatibility^20,21^. Therefore, we first selected poly(3,4-ethylenedioxythiophene):polystyrene sulfonate (PEDOT:PSS), which yields a porous interface rich in charge storage and capable of lowering impedance relative to bare gold^22^ (**Fig. 1d** and **Supplementary Fig. 9**). To control film thickness, we opted for a 100-s potentiostatic electrodeposition, which uniformly coated the etched sites and produced an order-of-magnitude reduction in impedance (**Fig. 1e**, **Supplementary Figs. 10** and **11**). However, PEDOT:PSS interfaces can delaminate during long-term exposure to biological environments^23^. Thus, we designed a surface-functionalization strategy using 11-mercapto-1-undecanol (MUD) followed by glycidyloxypropyltrimethoxysilane (GOPS) to promote covalent bonding between the conductive polymer and the gold surface (**Supplementary Fig. 12**). Contact-angle measurements confirmed the hydrophilic end-group deposition, Fourier transform infrared spectra (FTIR) identified epoxide functionality, and Raman spectra revealed a narrowed full width at half maximum indicative of increased PEDOT chain ordering^24^ (**Supplementary Figs. 13** and **14**). Strikingly, 30-day aging studies performed at 37 °C showed that both functionalized and pristine PEDOT:PSS coatings maintained sub-kilohm impedance with no measurable degradation, and the polyimide encapsulation exhibited no visible delamination at etched boundaries (**Fig. 1e** and **Supplementary Figs. 15** to **17**). These results allowed us to prioritize pristine PEDOT:PSS for rapid manufacturing cycles.

Surgical sutures require high pliability and low memory to pass smoothly through tissue and maintain secure fixation^25^. NeuroWeaves were thus engineered with mechanical properties that enable comparable handling during stitch-based implantation. The total device thickness (180–270 µm) yielded sufficient flexibility to achieve radii of curvature as small as 8.9 mm, which corresponds to the simulated yield point of the polyimide encapsulation layer^26^ (**Fig. 1f**). The thread-based construction allows NeuroWeaves to conform naturally to the scalp surface while matching the profile of standard scalp sutures^27^ (**Fig. 1g**). Tensile testing of bundled threads showed that five gold filaments produced a strength of ∼1.3 N, sufficient to withstand insertion forces (**Supplementary Fig. 18**). Applying medical adhesive further helped prevent breakage at the needle–thread junction (**Supplementary Fig. 19**). Sutured NeuroWeaves on porcine skin generated interfacial tensions below 40 mN, similar to surgical sutures (**Fig. 1g** and **Supplementary Fig. 20**). This suggests minimal tissue disruption or discomfort during placement for clinical deployment.

To support continuous ambulatory recording, we designed a wearable wireless unit inspired by the form factor of the external processors for cochlear implants (**Fig. 1h**). The device weighed 18.25 g when positioned behind the ear, reducing mechanical load and supporting day-to-day wear (**Figs. 1h** and **i**). Swappable magnetic connectors allowed the batteries to support eight-hour recording sessions and be exchanged without interrupting data collection (**Fig. 1j**). The printed circuit-board (PCB) layout can be found in Supplementary Fig. 21. Recorded signals are streamed in real-time to a handheld tablet or computer interface for monitoring and analysis, and the NeuroWeaves can also connect to clinical touch-proof leads during supervised evaluations (**Supplementary Fig. 21**).

### Acute preclinical validation

Prior to preclinical validation, we assessed the biocompatibility of all device materials. In vitro cytotoxicity and biocompatibility assays confirmed no adverse cellular responses, and all materials met ISO 10993-5 cytotoxicity requirements^28^ (**Supplementary Figs. 22** and **23**). We next evaluated the robustness of our PEDOT:PSS contacts using a customized roughened probe, and impedances in saline remained sub-kΩ after 10 abrasive cycles (**Supplementary Figs. 24** and **25**). To simulate placement within the epidermis, we repeated this experiment in vivo and observed stable contact impedances after 30 positioning cycles, which is more than sufficient for clinical handling (**Supplementary Figs. 26** and **27**).

To test broadband activity and signal fidelity, we sutured a 12-cm NeuroWeaves device with a 1-mm contact into the epidermis near the right primary sensory cortex of an anesthetized rat. Epidural screws were implanted on the frontal bone to serve as control electrodes. Both systems were connected to a commercial tethered acquisition unit (Intan Technologies LLC) to validate recording performance in the same animal (n = 3) (**Fig. 2a**). NeuroWeaves exhibited in vivo impedances in the expected range (mean 3.5 kΩ, n = 2 contacts) (**Fig. 2b**). This is consistent with the ∼570-fold smaller geometric surface area of a single NeuroWeave contact compared to an epidural screw (∼0.03 mm² versus ∼17 mm²). In addition, the commercial tethered system has an amplifier input impedance of 13 MΩ, which remains substantially larger than the electrode impedance, thereby minimizing voltage attenuation at the electrode–amplifier interface. Baseline recordings from both electrodes showed similar waveform fluctuations in raw and band-pass filtered (1 – 100 Hz) traces, with slightly larger amplitudes observed in epidural screws due to their closer proximity to cortical tissue (**Fig. 2c**). The signal-to-noise ratio (SNR) of the epidural screws was comparable to that of the NeuroWeaves (0.95 versus 0.92). However, the root-mean-square (RMS) amplitude of the baseline signal over a 5 s duration was higher for the epidural screws (28.18 µV) compared to the NeuroWeaves (18.17 µV). In addition, the filtered baseline traces and corresponding power spectral density (PSD) analysis are provided in Supplementary Fig. 28 for comparison between the two electrode types.

**Fig. 2.**
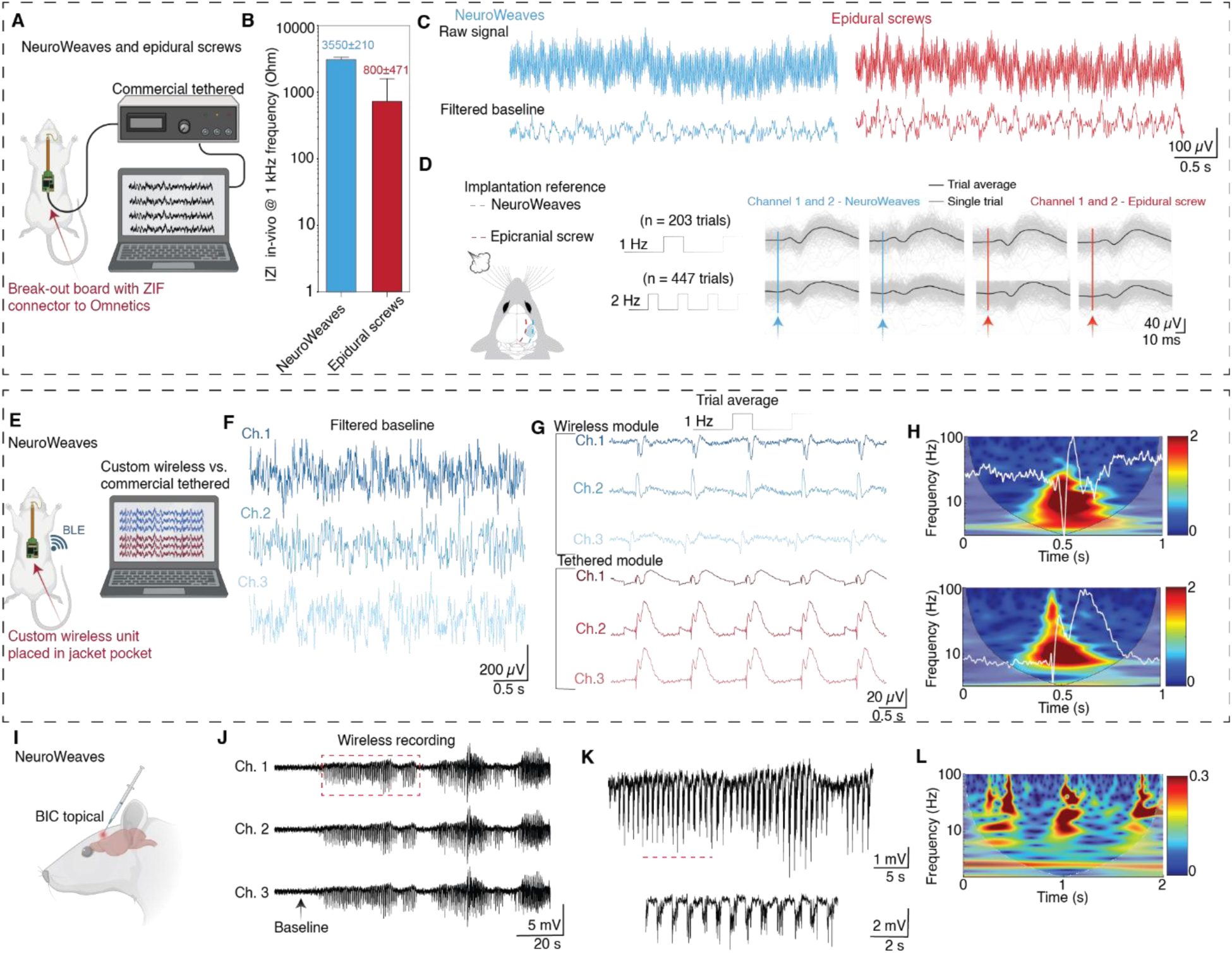
Acute preclinical validation. **a**, Schematic illustration of the electrode validation tests of NeuroWeaves compared to control epidural screws when tethered to a commercial system in the same animal (n = 3). **b**, In vivo impedances comparison between NeuroWeaves and epidural screws at 1 kHz frequency. **c**, Raw and filtered (1 – 100 Hz) baseline activity under anesthesia. **d**, Trial averaged responses (black lines) and single trials (grey) for all channels of NeuroWeaves and epidural screws at 1 Hz puff stimulation (n = 207 trials) and 2 Hz puff stimulation (n = 447 trials). **e**, Schematic illustration of the wireless validation tests of NeuroWeaves tethered to custom wireless board and NeuroWeaves tethered to commercial acquisition system (n = 3). **f**, Filtered (1 – 100 Hz) baseline of the three channels after transferring data to a handheld device under anesthesia. **g**, Comparison of the trial averaged whisker evoked potentials between custom wireless and commercial tethered system over a 5-second sliding window. **h**, Scalograms comparison of the custom wireless (top) and commercial tethered (bottom) of a 1-second trial averaged response. The tethered recordings exhibited broader stimulation-locked high-frequency transient components extending up to 100 Hz, whereas both systems showed peak spectral power concentrated within the lower-frequency range (∼5–20 Hz). **i**, Photographic image of the chemically induced seizure topically under anesthesia with NeuroWeaves device stitched near flap site and connected to the wireless board. **j**, Filtered (1 – 100 Hz) demonstrating evolution from baseline to seizure event for all channels. **k**, Zoomed-in window of the electrographic seizure from red dashed box in (j). Bottom trace shows a zoomed-in time window to demonstrate the rhythmic burst events from the top dashed line trace. **l**, Scalogram of a 2-second window shows peak power concentration of the seizure event in the ∼8–15 Hz and 20–80 Hz bands.

Whisker deflection in rats produces robust, time-locked somatosensory evoked potentials (SSEPs), providing a reliable model for assessing temporal fidelity of neural recordings^29,30^. We hypothesized that NeuroWeaves could capture these SSEPs transcutaneously because the skull-to-cortex separation is small in rats^31^. Air-puff stimuli were therefore delivered to all contralateral whiskers to generate a large, evoked response. Both electrode systems recorded clear SSEPs characterized by an initial positive deflection, a subsequent negative component, and a prominent late positive wave consistent with known late somatosensory activity^32^ (**Fig. 2d**). Notably, NeuroWeaves exhibited a mean latency of ∼11 ms compared to ∼15 ms for epidural screws, potentially reflecting differences in electrode location relative to the barrel cortex.

These location-dependent differences motivated us to test whether NeuroWeaves could resolve spatial variation in whisker-evoked responses. We next increased the number of NeuroWeaves contacts to four recordings channels to assess latency differences for spatial localization (**Supplementary Fig. 28**). The contacts were sutured in a loop spanning contralateral and ipsilateral positions. During contralateral stimulation near the whisker-barrel cortex, randomized trials showed a significant latency difference between contralateral and ipsilateral contacts (mean ∼11 ms versus ∼16 ms, p = 0.0026), whereas ipsilateral-only stimulation produced no significant difference (p = 0.0978) (**Supplementary Fig. 28**). Also, reproducibility across animals was confirmed by additional in vivo impedance, baseline, and whisker-evoked measurements (**Supplementary Fig. 29**).

After validating signal fidelity with the tethered system, we evaluated performance using our custom wireless platform that incorporates an input impedance of 1 GΩ (**Fig. 2e**). The wireless unit samples at 1,000 Hz and exhibits a −3 dB bandwidth at 342 Hz (**Supplementary Fig. 30**). Its signal-to-noise ratio is ∼15 dB at 1 µV peak-to-peak simulated waveform (**Supplementary Fig. 31**). This is sufficient to resolve the temporal dynamics of whisker-evoked responses. To benchmark wireless performance against the commercial tethered system, we implanted two NeuroWeaves devices in the same animal (n = 3) with three contacts each in parallel near the right somatosensory cortex—one connected to the Intan system and the other to the wireless unit. A handheld tablet placed inside the Faraday cage received the transmitted data, and benchtop testing demonstrated a mean packet-loss rate of ∼0.06% over a 2-hour recording within a 10-cm transmission distance. Filtered baseline activity acquired through the custom wireless system showed broadband neural dynamics across all channels under anesthesia (**Fig. 2f**), with SNR values ranging from 0.88 to 0.91. Here, the NeuroWeaves contacts were Here, the NeuroWeave contacts had an inter-electrode pitch of 1 cm and were sutured unilaterally. During puff stimulation, the custom wireless system captured clear, time-locked somatosensory-evoked potentials that closely matched recordings from the commercial tethered system when compared across sliding 5-second trial-averaged windows (**Fig. 2g**). Moreover, both systems produced comparable 1-second trial-averaged scalograms with similar temporal and spectral structure and peak power concentrated in the low-frequency band (∼5 – 20 Hz) (**Fig. 2h**). Additional quantitative time-frequency analysis was performed by separating the 1-second trial-averaged traces into frequency bands, including theta (4–8 Hz), alpha (8–12 Hz), beta (13–30 Hz), low gamma (30–70 Hz), and high gamma (70–100 Hz), and comparing the average power during the stimulation-locked evoked-response window against a pre-stimulation baseline window using 10log _10_(*P*_evoked_/*P*_baseline_). Such analysis demonstrated strong low-frequency enhancement in both systems, with theta and alpha increases of 20.52 dB and 17.56 dB in the commercial tethered recordings, and 14.82 dB and 18.29 dB in the custom recordings, respectively. The commercial tethered recordings also exhibited greater gamma-range enhancement, with low gamma and high gamma increases of 8.81 dB and 9.36 dB, respectively, compared to 2.31 dB and 5.27 dB in the wireless recordings. Hence, both systems captured robust stimulation-locked evoked activity, while the commercial tethered recordings preserved comparatively greater broadband high-frequency transient components than the custom wireless recordings. The source codes for the wireless recording system is publicly available at https://github.com/abdulhameedabdal/NeuroWeaves-codes. We repeated these comparative experiments across multiple animals and observed consistent agreement in signal amplitude, response latency, and spectral profiles between the two systems (**Supplementary Fig. 32**).

### Induction of seizure in preclinical models

Robust detection of pathological neural activity is essential for translational utility, so we evaluated NeuroWeaves during chemically induced seizures. The GABA_A_ receptor antagonist bicuculline was used to elicit focal cortical discharges^33^. A small scalp flap was elevated near the frontal bone, and a cranial opening was made to expose the dura. This allowed us to suture the NeuroWeaves in close proximity to the induction site. Bicuculline was dissolved in dimethyl sulfoxide (DMSO) to enhance membrane permeability^34,35^ and applied topically to the dura while recordings were acquired with the custom wireless system (**Fig. 2i**). Seizure onset occurred 23 minutes later, consistent with slow diffusion across the dura (**Fig. 2j**). The resulting activity consisted of burst–pause discharges with strong energy in the 8–15 Hz and 20–80 Hz bands that persisted for ∼160 seconds. These electrographic bursts were accompanied by whisker twitching temporally aligned with the activity, which is suggestive of early facial motor involvement and may represent an early clonic correlate^36^ (**Figs. 2j** to **k**).

To assess deeper limbic activity, we injected bicuculline into the amygdala while recording with the commercial tethered system. Band-pass (1 – 300 Hz) traces revealed spike discharges in two channels ∼12 seconds after induction, corresponding to contacts nearest the injection site (**Supplementary Fig. 33**). A ∼7 Hz rhythm emerged ∼40 seconds later with accompanying surface spikes (**Supplementary Figs. 34** and **35**), indicating cortical recruitment during propagation of the limbic seizure. These results show that NeuroWeaves capture seizure onset and propagation across brain regions essential for translational monitoring.

### Semi-chronic preclinical validation

Having established acute recording performance, we next evaluated NeuroWeaves in semi-chronic preparations to assess stability over extended timescales in freely moving rats (**Fig. 3a** to **c**). For these studies, the contact length was increased to 3 mm and the overall device length to 18 cm to improve mechanical stability, reduce the likelihood of contact delamination during extended implantation, and improve flexibility during freely moving conditions. To demonstrate material versatility, we fabricated NeuroWeaves using either FDA-cleared platinum nanorods (PtNR)^37^ or PEDOT:PSS coatings (**Fig. 3d** and **Supplementary Fig. 36**). Importantly, despite these dimensional modifications, the underlying device architecture, fabrication process, and recording principles remained unchanged. Devices were tunneled transcutaneously from the thoracic region for reference and ground channels, and the recording contacts were positioned over the cortex (**Supplementary Figs. 37**). Semi-chronic impedance measurements across materials and animals showed recording durations of ∼2 weeks (n = 3), with early termination primarily resulting from behavioral and packaging constraints, including jacket removal, device displacement, and connectorization failure, rather than material degradation.

**Fig. 3.**
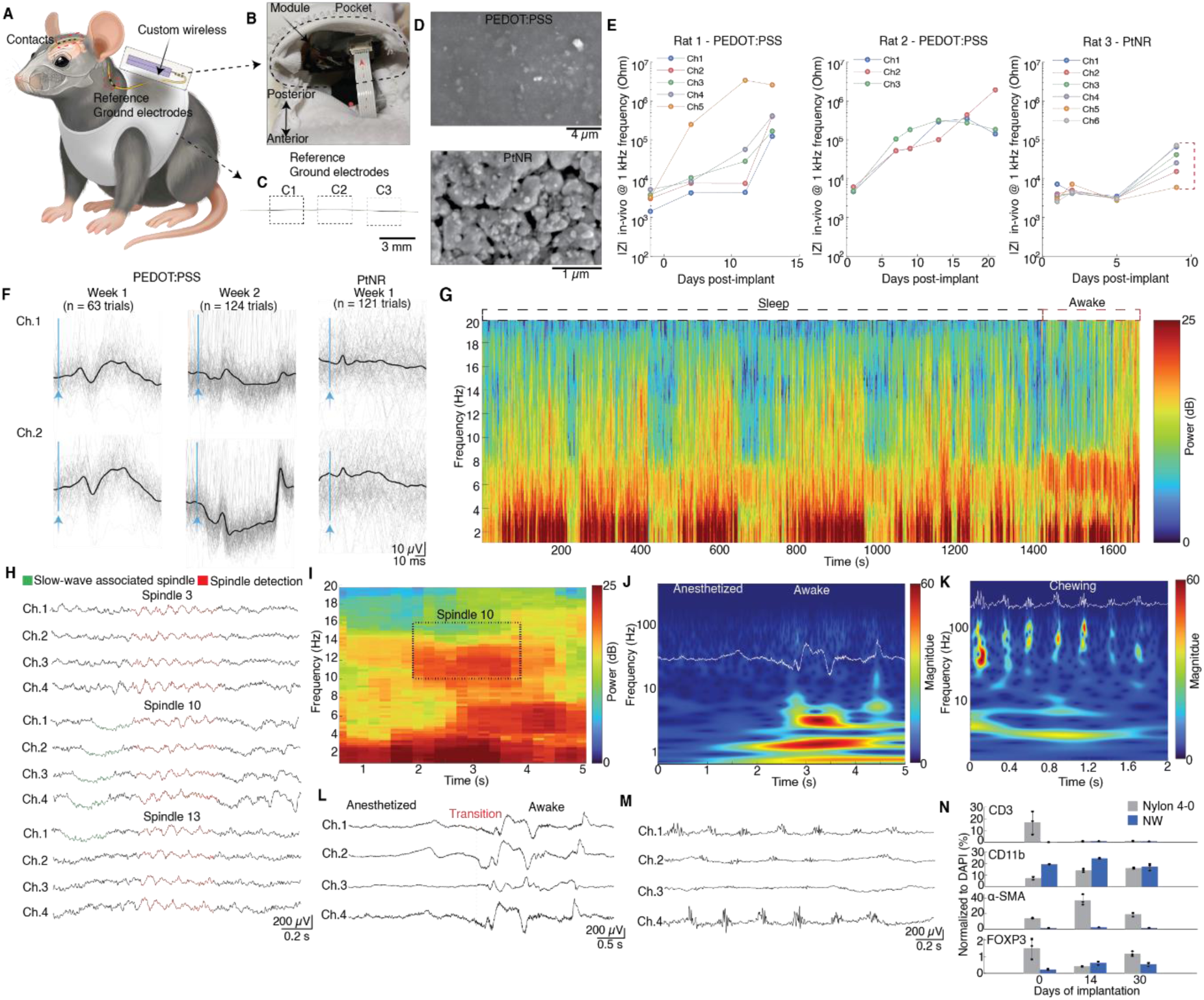
Semi-chronic preclinical validation. **a**, Schematic illustration of integrated wireless NeuroWeaves system in a freely-moving rat. **b,** Photographic image of the embedded wireless module in the pocket of the rat jacket. Red dashed line shows the connectorization of the break-out board from the NeuroWeaves connection-end to the wireless board via flexible cable connector. **c**, Photographic image showing the ground and reference electrodes with three 3-mm electrode widths design for ambulatory wireless demonstration. **d**, Scanning electron microscopy of the PEDOT:PSS contact (top) and FDA-cleared PtNR (bottom). **e**, In vivo impedances across three rats over extended periods of recordings and red-dashed line shows early termination of the PtNR due to NeuroWeaves relocation from excessive dynamic movement of the rat after 1-week post implantation. **f**, Trial averaged responses (black lines) and single trials (grey) for all materials over extended recording sessions when tethered to the commercial system for validation prior to connectorization of the wireless module for freely moving behavior recording sessions (n = 2). **g**, Multitaper spectrogram filtered showing an example of a sleep session with strong power concentrated in the low frequency bands for PtNR NeuroWeaves. Dashed lines show sleep and awake events. **h**, Spindles and slow-wave associated spindles across all recording channels when recording wirelessly. **i**, Inset of the multitaper spectrogram of Spindle 10 for channel 1 shows strong 10 – 14 Hz power concentration demonstrating spindle activity. **j**, Scalogram showing transition from anesthetized to awake state for channel 1 with trace overlay. **k**, Scalogram showing chewing events for channel 1 with trace overlay. **l**, Filtered (1 – 100 Hz) traces of all wirelessly recording channels for transition state from anesthesia to awake. **m**, Filtered (1 – 100 Hz) traces of all recording channels for chewing event. **n**, Biomarker studies for chronic periods between Nylon 4 – 0 surgical sutures and PEDOT:PSS NeuroWeaves showing minimal to no adverse events (n = 6).

To confirm electrode functionality over time, we performed whisker-puff validation under anesthesia for a mean of 10 days in PEDOT:PSS and PtNR NeuroWeaves (n = 2). Several recordings exhibited a polarity inversion of the evoked response, likely reflecting minor contact relocation (relative to the reference) associated with tissue healing or movement; sham controls confirmed the physiological origin of the responses (**Fig. 3f** and **Supplementary Fig. 38**). We then performed recordings in untethered rats and measured different behavioral activities (n = 2). For instance, we recorded ∼21 minutes of natural sleep and detected 16 spindles during this interval in PtNR NeuroWeaves (**Fig. 3g** and **Supplementary Figs. 39** to **41**). Notably, some spindles were preceded by slow-wave activity across multiple channels, others showed a preceding slow wave on only one channel, and some occurred without any preceding slow-wave activity (**Fig. 3h**). This variability is consistent with prior reports that spindle–slow-wave coupling is heterogeneous and that many—but not all—spindles are expressed in association with local slow waves or K-complex–like events^38^. These spindles exhibited strong 10 – 14 Hz activity, in agreement with characteristic rat spindle frequencies (10 – 16 Hz)^39^. To further illustrate freely moving behavior, NeuroWeaves recordings captured distinct activity patterns during emergence from anesthesia, spontaneous movements, and chewing artifacts (**Figs. 3j** to **M** and **Supplementary Movie 1**). Finally, histological analysis of CD3, CD11b, α-SMA, and FOXP3 showed minimal T-cell infiltration, limited myeloid recruitment, low fibrotic response, and stable regulatory T-cell activity over 30 days (**Fig. 3n**, **Supplementary Figs. 42** and **43**). All preclinical models, including experimental tasks, are summarized in **Table 4**.

### Human clinical validation

Translational feasibility requires adherence to clinical sterilization standards, and our fully assembled NeuroWeaves—mounted on clinical connectors—were compatible with conventional sterilization procedures^40^ (**Supplementary Fig. 44**). We next conducted a pilot clinical evaluation (n = 2) in a healthy participant. For these studies, devices were 18 cm in length with 3-mm contact windows, and surgical skin markers were used to guide suturing (**Supplementary Fig. 45**). As an initial demonstration, PEDOT:PSS NeuroWeaves were positioned over the occipital scalp and recorded a robust posterior dominant rhythm (PDR) that emerged after eye closure (**Supplementary Fig. 46**). In subsequent experiments, we maintained the NeuroWeaves near the occipital region and added two standard clinical electrodes at O2 and FP2 locations according to the international 10–20 system (**Fig. 4a**). Interfacial impedances were comparable across electrodes, with NeuroWeaves exhibiting slightly lower values than the clinical control (9.5 versus 10 kΩ) (**Fig. 4b**). Baseline EEG activity (1 – 100 Hz) showed NeuroWeaves with a narrower distribution of amplitudes, consistent with reduced baseline variability; Levene’s test confirmed unequal variances (p = 0.022) (**Supplementary Fig. 47**). Posterior dominant rhythm was reproducible across sessions, with electrodes near the occipital lobe capturing clear alpha-band activity (8 – 12 Hz) (**Fig. 4c**). These changes were corroborated by a mean power shift across electrodes between eyes-open and eyes-closed states (**Fig. 4d**) and by a statistically significant increase in alpha power for NeuroWeaves between open and closed conditions (p = 0.0002) (**Fig. 4e**).

**Fig. 4.**
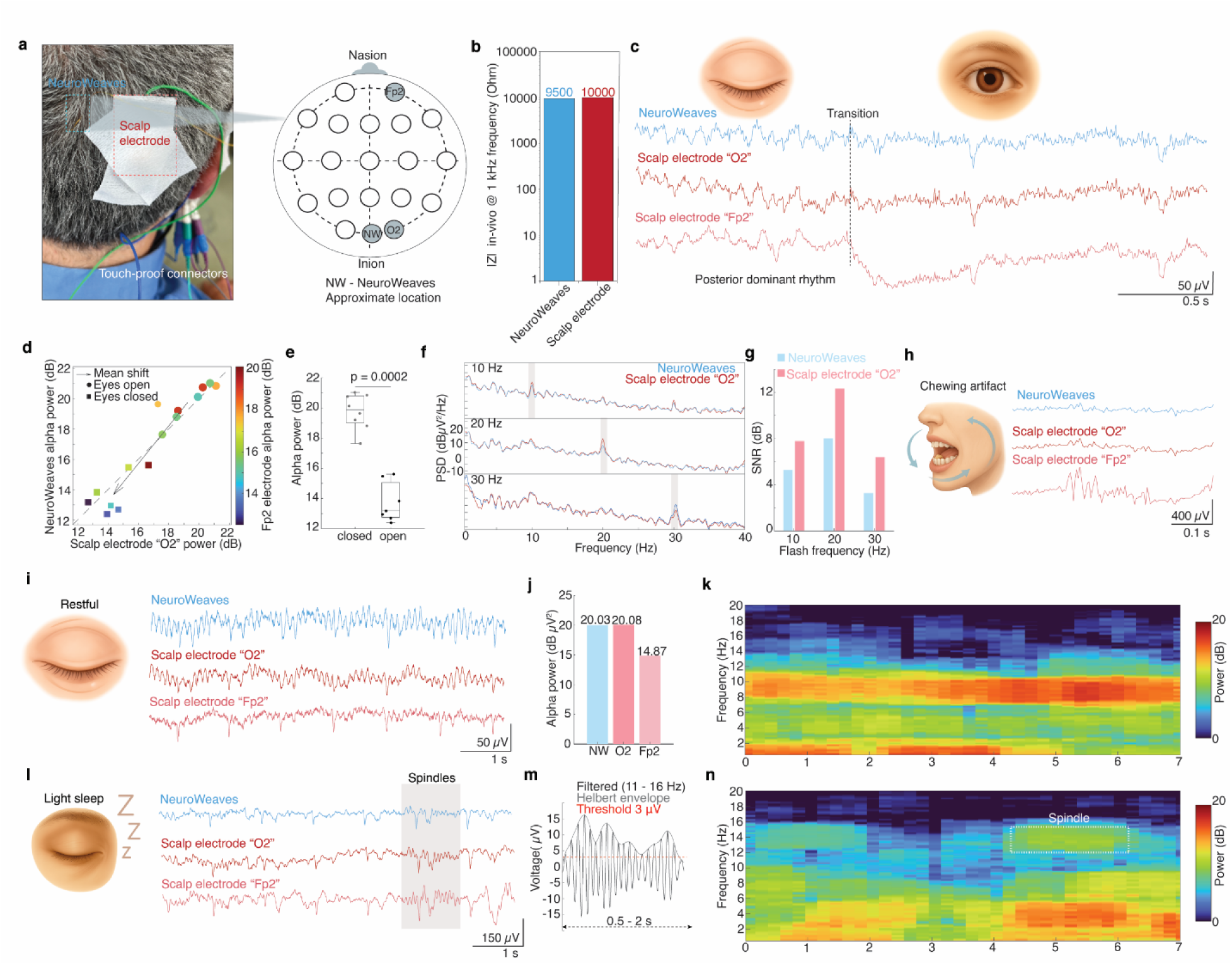
Human clinical validation. **a**, Photographic image showing NeuroWeaves implanted near the occipital region with clinical electrodes acting as control (left) (n = 2 trials). Approximate location of the NeuroWeaves according to the 10-20 International system (right). **b**, In vivo impedance comparison between NeuroWeaves channel versus clinical O2 electrode. **c**, Filtered traces (1 – 100 Hz) reveal transition state between eyes open and eyes closed for all channels showing strong PDR response for NeuroWeaves and O2 channels. **d**, Alpha power of all channels shows a strong convergence in the y = x line segment with a mean shift of power from eyes closed to eyes open. **e**, Windowed alpha power was greater during eyes-closed than eyes-open, confirmed by a permutation test (p = 0.0002). **f**, Power spectral densities across 10 – 30 Hz flash frequencies show clear peaks of the NeuroWeaves and scalp O2 electrode demonstrating driving photic response in a healthy subject. **g**, Comparable SNR between NeuroWeaves and O2 clinical electrode across flash frequencies. **h**, Filtered (1 – 100 Hz) traces of all channels during chewing event showing strong spikes for the FP2 clinical channel due to close proximity of the temporalis muscle. **i**, Restful awake event shows strong PDR for the filtered traces for the NeuroWeaves and O2 electrode. **j**, Alpha power distribution confirming strong PDR for the channels near the occipital scalp region. **k**, Multitaper spectrogram show power concentration in the ∼8 – 12 Hz during awake closed eyes. **l**, Detected spindle event during NREM showing waxing-waning morphology across recording channels. **m**, An example of a filtered trace (11 – 16 Hz) showing detected spindle when applying algorithm. **n**, Inset of the multitaper spectrogram show power concentration in the 11– 16 Hz for the spindle event.

Photic stimulation is routinely used to probe visual cortex responsiveness and to screen for photosensitivity^41^. In healthy individuals, EEG activity typically results in photic driving effect which represents repetitive visual evoked potentials^42^. We hypothesized that NeuroWeaves and the clinical O2 electrode, owing to their proximity to the visual cortex, would capture stimulus-locked responses to photic stimulation. Power spectral densities revealed clear peaks at the stimulation frequencies with substantial overlap between NeuroWeaves and O2 (**Fig. 4f**). Signal-to-noise ratios were comparable between the clinical electrode and NeuroWeaves across stimulation frequencies of 10, 20, and 30 Hz, with the clinical electrode exhibiting SNR values of 7.77 dB, 12.32 dB, and 6.38 dB, respectively, compared to 5.27 dB, 8.01 dB, and 3.28 dB for NeuroWeaves (**Fig. 4g**). The slightly lower NeuroWeaves power likely reflects small differences in electrode position relative to the underlying cortex. To assess sensitivity to movement-related artifacts, we induced chewing and observed the largest muscle activity in the clinical FP2 channel, which lies closest to the temporalis muscles (**Fig. 4h** and **Supplementary Fig. 48**).

Finally, to assess comfort and signal fidelity in a clinically relevant context, we conducted a short sleep study (**Supplementary Fig. 49**). During relaxed wakefulness, NeuroWeaves recorded clear alpha oscillations (**Figs. 4i** to **k**). As the subject transitioned into non–rapid eye movement (NREM) sleep, delta power gradually increased, and we detected spindle events of at least 500 ms duration with waxing–waning morphology and spectral content within the conventional spindle band (11 –16 Hz)^43^ (**Figs. 4l** to **n**). Post-explant inspection showed minimal to no visible damage to the recording contacts (**Supplementary Figs. 50** and **51**), supporting the mechanical robustness of the device during clinical handling.

## Discussion

The results presented here demonstrate that suturable thread-based neural interfaces can extend electrophysiological monitoring into a regime that has traditionally required rigid electrodes or surgical implantation. NeuroWeaves integrate materials engineering, device architecture, and clinical workflow considerations to overcome long-standing issues of comfort, invasiveness, and spatial constraint. By combining hair-thin gold–polyimide filaments with low-impedance conductive coatings and a lightweight custom wireless platform, the system achieves stable recordings across acute, semi-chronic, and human pilot studies. These capabilities suggest that epidermal suturing—long used in routine medical practice—can serve as a viable route for transcutaneous neural access without the burden of gels, adhesives, or surgical exposure.

Several observations in this work open new opportunities for future exploration. The ability to resolve whisker-evoked responses, seizure dynamics, sleep rhythms, and photic entrainment using the same stitched interface suggests that the spatial reach of volume conduction may be more permissive. Differences in latency across sutured positions also hint at the potential for simple stitch patterns to extract spatial information, even without penetrating or intracranial placement. Moreover, our semi-chronic results indicate that thread-based contacts can remain stable in freely moving animals despite tissue motion, healing, and behavioral perturbations, an encouraging sign for long-term human use.

Human pilot data further revealed that NeuroWeaves can reproduce typical physiological signatures—including posterior dominant rhythm, sleep transitions, and photic responses –at a level comparable to clinical electrodes. This raises the possibility that suturable interfaces could complement or, in selected contexts, substitute for conventional surface EEG systems. The simplicity of implantation also suggests potential for decentralized or home-based monitoring, particularly in settings where access to prolonged EEG remains limited.

Despite these promising findings, several practical challenges remain. Although semi-chronic recordings demonstrated stable performance over approximately two weeks, the achievable duration was ultimately constrained by interactions with the externalized system, including jacket removal, displacement, and connectorization failure. These observations suggest that future improvements in packaging, wireless miniaturization, and implantation strategies will be important for extending recording duration and supporting scalable clinical translation.

## Outlook

Critically, the suturable mechanical fixation and direct epidermal interface of NeuroWeaves provide inherent stability analogous to surgical sutures, supporting continuous recordings over days to weeks without gel degradation or adhesive failure, and positioning this platform for truly extended ambulatory use.

Finally, we emphasize that the current implementation represents only the first iteration of this modality. Future versions could incorporate higher-density layouts, multimodal sensing, or closed-loop capabilities while retaining compatibility with suture-based deployment. Such advances, combined with the clinical usability demonstrated here, may help establish a new class of minimally invasive neural interfaces that bridge the gap between laboratory-grade electrophysiology and everyday neurotechnology for at-home use.

## Materials and Methods

### Fabrication of NeuroWeaves

#### (1) Photolithography steps

NeuroWeaves were fabricated using a trench-assisted wire transfer process adapted for long, insulated gold microwires (California Fine Wire Co.). SU-8 2005 (Kayaku Advanced Materials Inc) was spin-coated onto a silicon wafer and patterned by photolithography to form ∼65 µm-wide trenches that guided wire placement. Gold wires (8 cm length) were cut from a spool, straightened using isopropyl alcohol and gentle rolling, and manually positioned into the SU-8 trenches.

A separate glass wafer was coated with a thick layer of positive photoresist (AZ 12XT-20PL-10, MicroChemicals; 12–14 μm) and partially cured to form an adhesive surface. This semi-cured film was then brought into contact with the silicon wafer, enabling transfer of the gold wires from the SU-8 trenches onto the glass substrate during heating. Following transfer, the wafer was fully cured on a hot plate at 110 °C to stabilize the embedded wires. A second layer of positive photoresist was subsequently spin-coated and cured to encapsulate the wires and remove residual solvent.

Photolithography using a Karl Suss MA6 mask aligner and mylar photomasks (FineLine Imaging) defined openings along the wire length corresponding to electrical contacts and interconnects. A brief flood exposure was applied prior to etching to suppress foaming of the positive thick photoresist. Selective removal of the polyimide insulation was then performed using a mixed oxygen and tetrafluoromethane plasma for 30 – 40 minutes in 5-minute intervals by inductively coupled plasma reactive ion etching (ICP-RIE) at 100W-10W power ratio while maintaining temperature control at 20° C (Trion Minilock Phantom III ICP, Trion Technology). This cooling setup within 5-minute intervals minimized microcracking of the thick photoresist, and produced exposed regions for electrical interfacing and recording channels.

#### (2) Hard-mask process

A thermal-release–tape (TRT)–assisted process was used to microfabricate long insulated gold threads with patterned exposed regions. Thermal release tape (Semiconductor Equipment Corp.) was laser-cut using a CO₂ laser to define the wire placement layout and laminated onto 6-in or 8-in silicon wafers. Gold wires were cut to the required lengths, straightened using isopropyl alcohol and gentle rolling, and placed onto the predefined TRT regions. Both wafer formats were used: the 6-in TRT layout incorporated serpentine designs accommodating thread lengths up to ∼12 cm and provided four TRT lanes per wafer, each carrying two wires; the 8-in format used straight-line patterns supporting 16–18 cm threads and offered up to ten TRT lanes per wafer, each holding four wires, enabling significantly higher throughput.

A custom aluminum shadow mask (∼1 mm thick) was then aligned on top of the wafer to expose the intended contact and interconnect sites. Selective removal of the polyimide insulation was performed by dry etching in a mixed oxygen and tetrafluoromethane plasma for 30 – 40 minutes at 200 W RIE (Oxford Plasmalab 80+, Oxford Instruments). After etching, the wafer was placed on a 120 °C hot plate for at least two minutes to release the wires from the TRT. Retrieved wires were prepared for electrochemical testing by attaching copper leads with silver conductive paint (PELCO Conductive Silver Paint, Ted Pella Inc.) at the connection sites for impedance spectroscopy in phosphate-buffered saline (silver-silver chloride (Ag/AgCl) reference, platinum (Pt) counter electrode).

### Integration and Connectorization

After deposition of the conductive material, individual threads were mounted onto a custom mechanical assembly tool and twisted to form the NeuroWeaves bundle. The distal tip of the NeuroWeave, positioned at least 1 cm from the first electrode contact, was then threaded through a medical needle (Richard-Allan®) to create the attachment knot. To mitigate mechanical stress at the needle–NeuroWeaves interface, a biocompatible medical adhesive (215-CTH-LV-UR-SC/10SYMR, Dymax®) was applied to the knot region.

For connectorization, at least 0.5 cm of insulation was removed from the end of each NeuroWeaves bundle to allow attachment to an interconnector or breakout interface forthe custom wireless and commercial tethered systems.. These exposed endpoints were soldered using low-temperature solder paste (SMDLTLFP10T5, CHIPQUICK). When touch-proof clinical leads were used, the exposed NeuroWeaves wires were soldered directly to the lead terminals, and a rigid support structure was added to minimize mechanical load on the threads.

### Porcine skin incision experiments

Porcine test material was purchased from Stellen Medical, LLC (Cat. No. I-188) to perform the incision experiments. A mechanical testing apparatus (Mark-10) was used to evaluate knot strength and interfacial tension (passing force) for NeuroWeaves and Nylon 4-0 sutures (ETHILON®)) on ∼1 mm thick porcine skin. For knot strength measurements, the needle was inserted halfway through the skin to allow the probe to grasp the needle body (**Supplementary Fig. 20a**), and the knot strength was defined as the tension required for the knot to pass through the tissue. These tests were performed using a 100 N force gauge.

For interfacial tension experiments, the needle was fully passed through the skin so that only the tension transmitted through the suture was measured (**Supplementary Fig. 20b**). A slight increase in interfacial tension was observed, likely due to gradual drying of the porcine tissue. These measurements were acquired using a 0.5 N force gauge.

### Surgical procedures of preclinical cases

All animal experiments were approved by the University of California San Diego (UCSD) Institutional Animal Care and Use Committee under protocol S16020. Animals were initially induced with isoflurane anesthesia (4 – 5%), and all surgical procedures were conducted under maintained isoflurane (2 – 3%) delivered through a precision vaporizer and scavenged with charcoal filters. Anesthetic depth was continuously monitored by tracking heart rate (MouseSTAT® Jr.), respiratory rate, and responsiveness to toe pinch, while body temperature was maintained using a heated waterbed. Before recording, each animal was transitioned from isoflurane to a ketamine/xylazine cocktail (80/20), and data collection commenced only after full withdrawal from isoflurane. Subdermal needle electrodes were used for ground and reference and were placed along the scalp midline, with minor adjustments made to minimize ECG artifacts.

#### (1) Acute recording cases

For acute experiments, NeuroWeaves were sutured subdermally on the scalp above the right hemisphere, centered approximately over the somatosensory cortex, such that the number of skin insertions matched the number of device contacts. In some experiments, stainless-steel screws (1/8" EEG Screw, Pinnacle Technology De, LLC) were implanted alongside the NeuroWeaves to enable recording comparisons. These screws were positioned along the anterior–posterior axis of the frontal and parietal bones on the right hemisphere with an approximate pitch of 0.5 cm. To evoke sensory activity, air-puff whisker stimulation (PV830 Pneumatic PicoPump, WPI) was delivered through a microcapillary tube to move the whiskers on the contralateral side at 40 psi.

All NeuroWeaves used in these studies had a contact width of 10 µm, an inter-electrode pitch of 1 cm, and a total device length of 12 cm. For electrode comparison tests (n = 3), recordings were acquired using a commercial tethered system (Intan Technologies LLC). For wireless benchmarking (n = 3), NeuroWeaves were recorded using both the commercial tethered system and our custom wireless platform for direct comparison. **Table 4** summarizes all experiments performed in the acute cohort.

#### (2) Induction of seizures

To induce seizures, a 3 × 3 mm craniotomy was created over the somatosensory cortex using a surgical drill. Bicuculline (5 mM in 10 μL DMSO) was administered either by topical application onto the exposed cortex or by direct injection into the amygdala (−2.8 mm anteroposterior, 5 mm mediolateral, 8.8 mm dorsoventral) using a 1 μL Hamilton syringe mounted on a stereotaxic holder. **Table 4** summarizes all experiments performed for seizures.

#### (3) Semi-chronic cases

For chronic experiments, a fully implanted approach was used to prevent animals from scratching or dislodging the NeuroWeaves. A 1 cm scalp incision was made, followed by gentle tissue scraping to expose a clean, dry bone surface. A secondary incision was created on the dorsal thoracic region approximately 5 cm caudal to the head. The NeuroWeaves were then tunneled subcutaneously from the back incision to the head using fine surgical forceps. The distal tip of the NeuroWeave was secured to the skull using a UV-curable polymer (Tetric Evoflow Syringe Trans, 2 g). The device breakout connector was permanently housed within the animal jacket (Lomir Biomedical Inc.). Prior to wireless recording sessions, NeuroWeaves were connected to the commercial tethered acquisition system to measure electrochemical impedances. During recordings, a wireless system with a battery was attached to the breakout connector and placed securely inside the jacket pocket and subsequently removed after each session. On separate days, whisker-barrel stimulation was performed to evaluate recording performance across the week. **Table 4** summarizes all experiments performed in the chronic cohort.

### Clinical procedure

An epilepsy monitoring unit technologist assisted in the preparation and adhesion of the scalp EEG. Prior to the recording, the human subject’s scalp was prepared at the intended scalp EEG adhesion and NeuroWeave suture sites using alcohol wipes and an abrasive gel, per the institution’s clinical standards, to minimize impedance and artifacts. The FP2 and O2 scalp EEG channels were placed using the international 10-20 system for EEG placement^44^, which correspond to approximately the right frontal and right occipital regions, respectively. These regions were chosen to gather physiologic signals, such as a posterior dominant rhythm (from the O2 channel) and eye blinking artifacts (from the FP2 channel). An electrode was placed on the chest to serve a reference. Following sterilization, the NeuroWeaves electrode was sutured in place, using a needle driver, next to the O2 electrode by a physician to primarily detect the posterior dominant rhythm.

A 1-hour recording, similar in duration to the usual routine EEG recordings performed at our institution, was performed and captured several states, including awake, drowsiness, and stage II sleep. An impedance check was conducted on the NeuroWeaves, and approximately 9.5kOhm was noted a few minutes after application, which was comparable to the EEG leads. During the awake state, normal physiologic data was captured, and the subject was asked to perform similar tasks to those that occur during a typical clinical routine EEG recording, including: opening and closing their eyes (to capture blinking and posterior dominant rhythm appearance), moving their head, and chewing. Additionally, photic stimulation was performed at increasing frequencies up to 30 Hz to capture photic driving, a typical physiologic response. Finally, the patient was allowed to rest for 20 minutes, and characteristic features of drowsiness and stage II sleep were seen.

After the recording was completed, the NeuroWeaves electrode and EEG electrodes were removed and the sites were cleaned with minimal bleeding noted. There was minimal discomfort during the NeuroWeaves suturing and none during removal, and there was no pain noted later that day or a week later; on a 1-week post-procedure check-in by a physician, there was no inflammation or infection noted at the site. All procedures and consent were conducted under the auspices of the local Institutional Review Board.

## Data Availability

All data obtained in this study are either presented in the paper and the Supplementary Materials or deposited in open database. Animal brain recording data could be accessed at DANDI (https://dandiarchive.org/), and the human brain recording data could be found in Data Archive BRAIN Initiative (DABI) (https://dabi.loni.usc.edu/).The manuscript includes all the data collected and analyzed in the study, and more information is included in the Supplementary Information.

## Code Availability

Custom MATLAB code (version R2021a) and analysis are available on GitHub (https://github.com/abdulhameedabdal/).

## Acknowledgments

We acknowledge insightful discussions with A. Bourhis of UC San Diego. This work was performed, in part, at the San Diego Nanotechnology Infrastructure (SDNI) of UCSD, a member of the National Nanotechnology Coordinated Infrastructure, which is supported by the NSF (grant ECCS1542148). Tissue Technology Shared Resource is supported by a National Cancer Institute Cancer Center Support Grant (CCSG Grant P30CA23100). Elements of Fig. 2 (A, E, I) were illustrated by BioRender.com

## Funding

A.A. acknowledges financial support from the Kuwait Foundation for the Advancement of Sciences (KFAS). R.B. acknowledges support by the Weizmann Institute of Science Women’s Postdoctoral Career Development Award. This was work was supported in part by the Air Force Office of Scientific Research (AFOSR) grant no. FA9550-19-1-0278 (to D.J.L.) and by the BRAIN® Initiative NIH grant UG3NS123723 (to S.A.D.).

## Author contributions

Conceptualization: A.A., H.K., D.J.L., S.S.C., S.A.D.

Methodology: A.A., H.K., F.K., P.H., B.F.C., P.S., G.C., J.C., A.N., S.K., S.J.E., R.V., M.H., T.R., J.W., R.B., K.S., T.W., J.J., D.J.L., S.S.C., S.A.D.

Investigation: A.A., H.K., D.J.L., P.H., S.S.C., S.A.D.

Visualization: A.A., H.K., F.K., P.H. B.F.C., S.A.D.

Funding acquisition: D.J.L., S.A.D.

Project administration: A.A., H.K., F.K., S.A.D.

Supervision: S.A.D.

Writing – original draft: A.A., S.A.D.

Writing – review & editing: A.A., H.K., P.H., D.J.L., S.S.C., S.A.D.

## Competing interests

The authors declare the following competing interests: S.A.D have shares in Cortical Science Inc. that licensed a patent disclosure related to this work. The other authors declare that they have no competing interests.

## Supplementary Information

Supplementary Discussions 1 – 10, reference, Figs. 1 – 51, Supplementary Movie 1, and Tables 1 – 4.

## Notes

### Clinical Trial

This was a a self-experimentation. All procedures and consent were conducted under the auspices of the local Institutional Review Board.

### Author Declarations

Ethics committee/IRB of Massachusetts General Hospital waived ethical approval for this work.

